# One minute Multiple Pupillary Frequency Tagging test to assess visual field defects

**DOI:** 10.1101/2022.01.24.22269632

**Authors:** Suzon Ajasse, Catherine Vignal-Clermont, Saddek Mohand-Saïd, Cecilia Coen, Carole Romand, Jean Lorenceau

## Abstract

Pupillary responses to light offer a convenient and objective way to quickly assess the functional health of the anterior afferent visual pathways. We here present a proof of concept of an innovative one minute pupillary test consisting in 9 visual sub-regions simultaneously modulated in luminance at 9 different temporal frequencies. The spectral power of the sustained pupillary responses evoked by this display over 45 seconds of passive fixation distinguishes patients with retinal and optic nerve diseases from healthy participants with remarkable sensitivity and specificity, at both global and local scales. Reliable and fast, this test could ease patient care and allow screening for, and following-up, chronic ophthalmic diseases whose prevalence worryingly increases worldwide.

## Introduction

The functional evaluation of vision in an important step in diagnosing and treating chronic ophthalmic diseases, such as glaucoma, diabetic retinopathy, AMD, etc., whose prevalence worryingly increases worldwide^1,2^. As a general rule, individuals consult only after they felt a visual loss, which often appears after structural damages occurred^3,4^. The functional evaluation of vision is performed at the clinic or at the hospital using subjective tests where individuals are asked to report on their visual percepts (for instance to assess visual acuity, or to perform Standard Automated Perimetry, SAP). Both the patients and the practitioners agree to consider that SAP is tedious and long^5^. In addition the results can be hampered by fluctuations of attention, eye-movements, fatigue, or stress. As a consequence SAP, performed in already overbooked ophthalmologic services, is unreliable^6^ and is only done once or twice a year, limiting its usefulness for following-up the evolution of the disease. Attempts to shorten the SAP exam have been made^7^, but these do not avoid some of the above mentioned issues.

Other studies aim at assessing visual loss by analyzing the pupillary response to light stimuli. Most often, these studies focus on parameters of the pupil light reflex (PLR) elicited by brief flashes of light^8^. Of interest are the pupil constriction latency, the constriction speed, the maximum constriction amplitude and its latency, as well as the time-course of the post-illumination response (PIPR) characterizing the return to base-line pupil diameter. The stimuli used to elicit a PLR can be full-field or focal localized flashes projected in different regions of the visual field^9–11^. Some studies use different stimuli (e.g. ramping full-field stimuli^12^), but the general outcome is that pupillary activity is perturbed in a number of ophthalmic diseases (Glaucoma, diabetic retinopathies, optic neuritis, etc.^13–15^), and can therefore be used to probe functional deficits. One consequence of these experimental studies is the search for convenient pupillary tests that can be used in routine clinical assessment to complete or replace the current examination of diseases related functional losses. As a matter of fact, in the context of the worldwide increasing prevalence of ophthalmic diseases, using pupillary activity to probe functional deficits has several advantages: pupillary responses are reflexive physiological objective signals that can be measured non-invasively with little expertise and resources in a short amount of time, dispending individuals to give a subjective, criterion dependent, response. Moreover, they are correlated to structural deficits (RNFL, GCC) measured by Optical Coherence Tomography (OCT)^12,16,17^. Pupillary activity can thus be used for screening populations at risks, which is otherwise difficult with current devices only available in clinics or hospitals, or for following-up already identified patients^18^.

One extensively documented method, multifocal Objective Pupillographic Perimetry (mfPOP) has been developed by Maddess and colleagues^10,10,19,20^. Although different versions were tested in different studies on different diseases, the core principle of mfPOP relies on eliciting many focal PLRs throughout the visual field, after dark adaptation. mfPOP employs brief high luminance flashes to isolate successive PLRs and efforts to improve signal-to-noise ratio and to shorten the examination time (from 7 minutes to 80 sec.) were made^21^. The many different reports of this group indicates that mfPOP is a powerful and sensitive method to detect and follow-up several neuropathies and retinopathies.

## Multiple Frequency Pupillary Tagging

We here present a proof-of concept of an alternative method, Multiple Frequency Pupillary Tagging (MFPT) to assessing a “pupillary field” at once in a short amount of time (<1 min per eye). Our method uses 9 simultaneously displayed visual sectors, each modulated in luminance at a specific temporal frequency (Frequencies of Interest, FOIs, from 1 to 3.5 Hz, Figure 1A). The rationale is that during passive fixation, each sector contributes to the overall pupillary response *at its corresponding frequency* (Frequency Tagging). In healthy individuals, the contribution of each modulated sector should be identifiable in the power spectrum of the sustained pupillary responses evoked by during fixation of our multipartite stimulus (Figure 1B). In individuals with a neuropathy or retinopathy, the pupillary power should be less –or null-in defective regions (scotoma), thus reflecting functional losses proper to the individual and the disease at stake.

**Figure 1:**
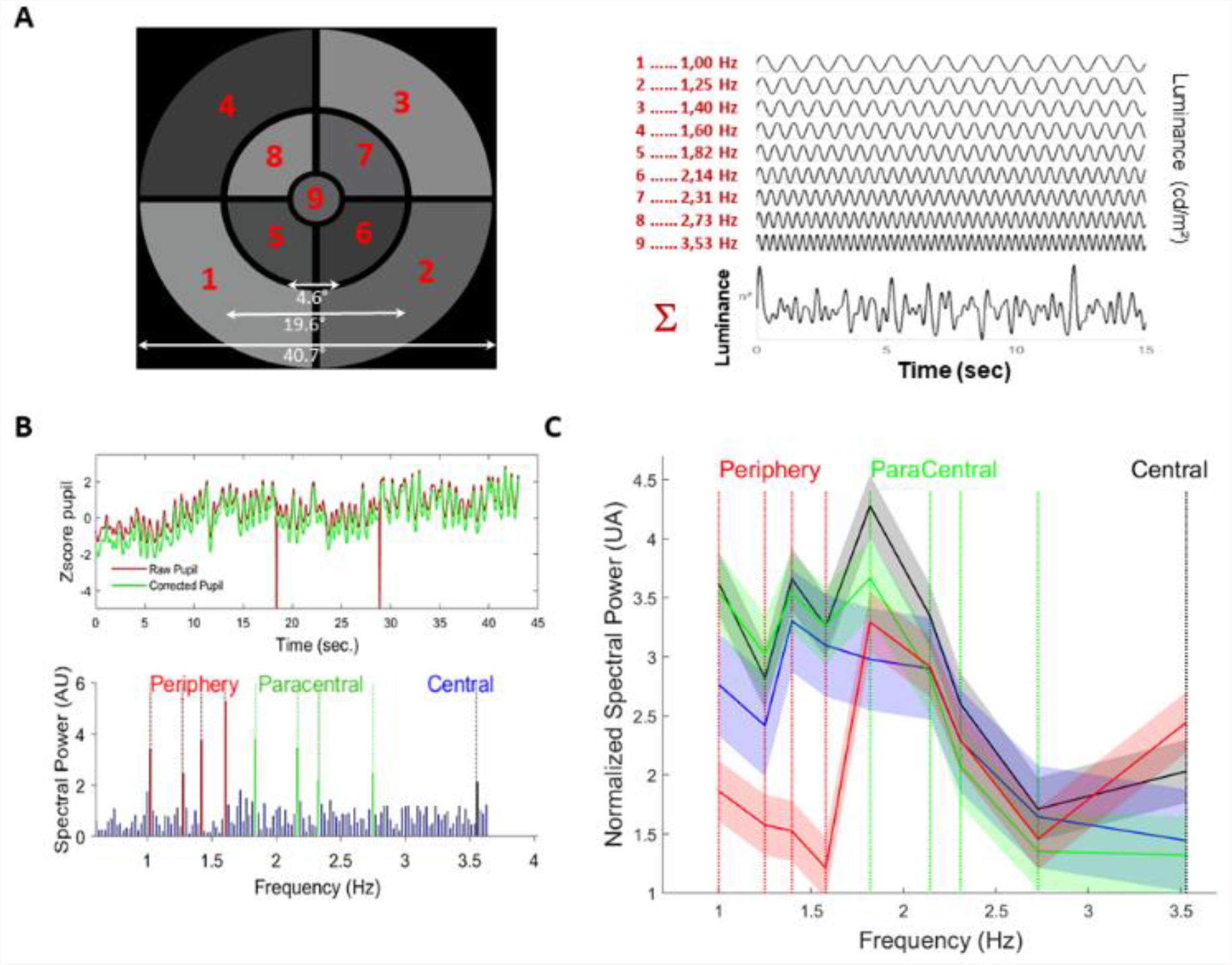
**A**. Display used in the study. *Left*: Nine sectors covering 40° of visual angle are sinusoidally modulated in luminance at 9 different temporal frequencies (*Right*). **B**. *Top*: Example of a raw and blink corrected pupil recording during a 45 seconds trial for a healthy participant. *Bottom*: Normalized spectral power distribution in the range 0.2 – 4 Hz, showing peaks at each of the 9 FOIs tagging each of the 9 sectors. **C**. Averaged spectral power and 95% confidence intervals at FOIs for the 4 groups of participants (Black: Healthy Participants, HP; Red: Retinitis Pigmentosa, RP; Green: Stargardt Disease, SD; Blue: Leber Hereditary Optic Neuropathy, LHON).

We tested this approach in a clinical study involving patients with Retinitis Pigmentosa (RP, n=14), Stargardt disease (SD, n=14), Leber hereditary optic neuropathy (LHON, n=9) and compared their pupillary fields with those of healthy individuals (HP, n=14). For this proof-of-concept study, our aim was to evaluate the feasibility and the acceptability of the test, and to determine its sensitivity and specificity with well characterized, although rare, diseases. The choice of these pathologies was motivated by the fact that these patients are relatively young adults, less likely to have comorbid pathologies or to take medications that could modulate the pupillary responses.

Before the main study, we conducted extensive preliminary experiments on healthy individuals to determine the optimal spatio-temporal configuration, the minimum amplitude of luminance modulations and the minimum test duration that still evoked reliable pupillary spectral power at FOIs. These preliminary experiments indicated that up to 9 TMFs could be mixed while still getting well identified components in the Fourier power spectrum of the pupillary responses. The minimum duration was evaluated to 45 seconds of stimulation with luminance modulations of moderate amplitude^22^.

## Results

In healthy participants (HP, n=14), the spectral pupillary power at FOIs (SPPf) revealed peaks at each of the 9 FOIs for each eye, indicating that each sub-region contributed to the overall pupillary response (Figure 1B). In patients diagnosed with Retinitis Pigmentosa (RP, n=14), Leber Hereditary Optic Neuropathy (LHON, n=9), and Stargardt disease (SD, n=14), the spectral power exhibited disease dependent distortions at FOIs, indexing specific sub-regions (Figure 1C), in line with the visual defects proper to these diseases (see Supplementary Table 1 describing the participants’ profiles).

As pupillary responses are larger for nasal as compared to temporal stimulations^23^, we first determined whether MFPT discriminate the left and right eye. AUC of ROC curves were high for HP (AUC=0.97), SD (AUC=0.95) and LHON (AUC=0.97), showing that the left and right eyes indeed have distinct spectral signatures for these subjects (Supplementary Table 2, Supplementary Figure 1). AUC of ROC was smaller for RP patients (AUC=0.69), in line with the defective peripheral vision of these patients that entails diminished contributions of nasal and temporal fields to pupillary responses. Importantly, these results further indicate that the SPPf is linked to sub-regions and not to particular temporal frequencies, which could have been a potential confound if temporal frequencies *per se* would elicit very different pupillary responses.

We then used the distribution of SPPf of each eye to discriminate healthy participants from patients. The AUC of ROC curves, sensitivity and specificity were remarkably high for each pathology and each eye (Table 1). In addition, we computed AUC of ROC separately for each FOI tagged stimulus sector, and could reliably identify defective sub-regions for each pathology (Figure 2).

**Table 1:**
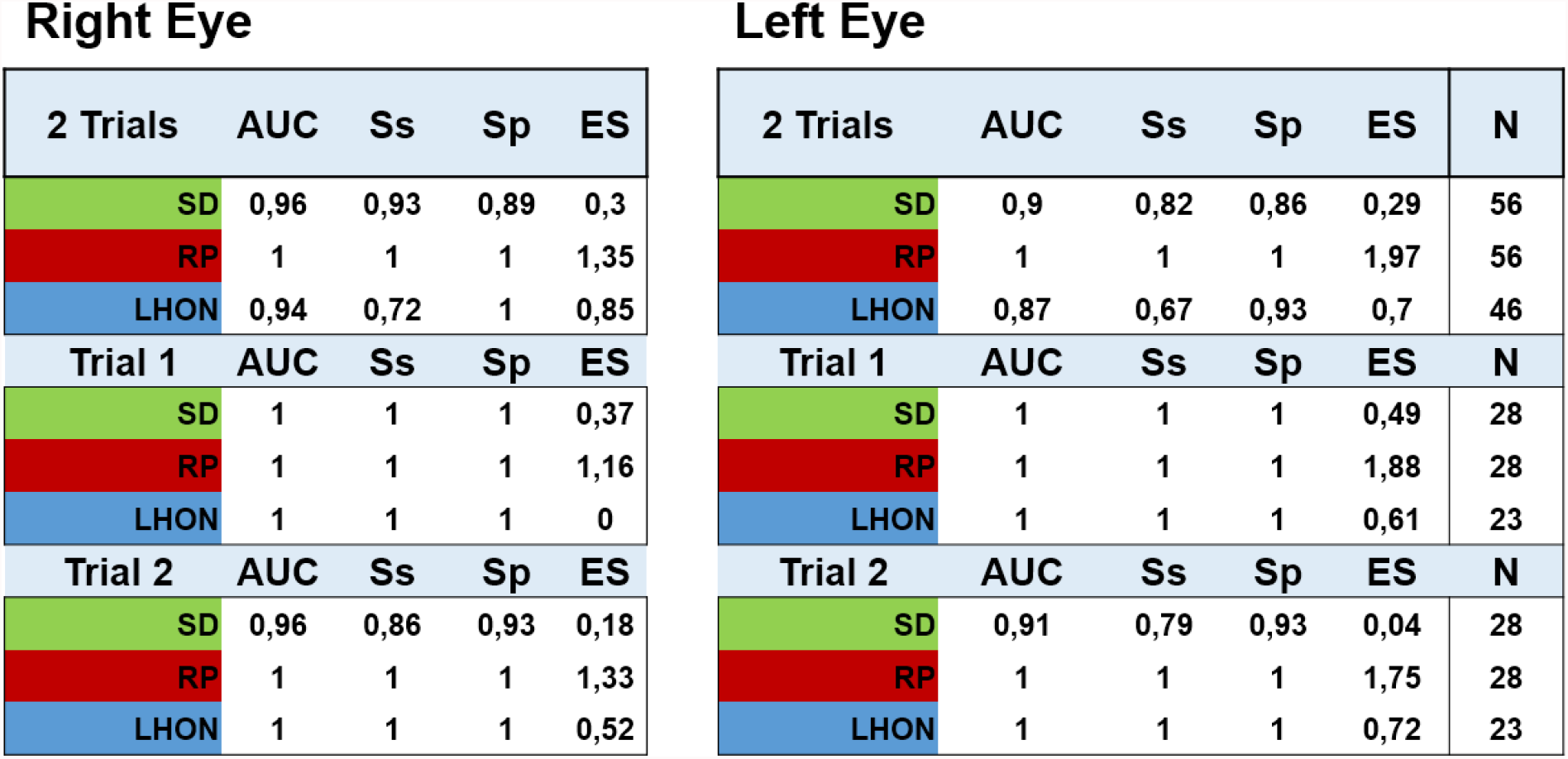
AUC of ROC, Sensitivity, Specificity and Effect Size (ES: Hedges m) for RP, SD & LHON patients computed against healthy participants using the spectral power of the 9 FOIs of 2 trials, and for each trial separately, for the Right and Left eyes. The number of observations is indicated in the last column.

Right Eye
| 2 Trials | AUC | Ss | Sp | ES |
| --- | --- | --- | --- | --- |
| SD | 0,96 | 0,93 | 0,89 | 0,3 |
| RP | 1 | 1 | 1 | 1,35 |
| LHON | 0,94 | 0,72 | 1 | 0,85 |
| Trial 1 | AUC | Ss | Sp | ES |
| SD | 1 | 1 | 1 | 0,37 |
| RP | 1 | 1 | 1 | 1,16 |
| LHON | 1 | 1 | 1 | 0 |
| Trial 2 | AUC | Ss | Sp | ES |
| SD | 0,96 | 0,86 | 0,93 | 0,18 |
| RP | 1 | 1 | 1 | 1,33 |
| LHON | 1 | 1 | 1 | 0,52 |

**Table 1:** AUC of ROC, Sensitivity, Specificity and Effect Size (ES: Hedges m) for RP, SD & LHON patients computed against healthy participants using the spectral power of the 9 FOIs of 2 trials, and for each trial separately, for the Right and Left eyes.
| 2 Trials | AUC | Ss | Sp | ES | N |
| --- | --- | --- | --- | --- | --- |
| SD | 0,9 | 0,82 | 0,86 | 0,29 | 56 |
| RP | 1 | 1 | 1 | 1,97 | 56 |
| LHON | 0,87 | 0,67 | 0,93 | 0,7 | 46 |
| Trial 1 | AUC | Ss | Sp | ES | N |
| SD | 1 | 1 | 1 | 0,49 | 28 |
| RP | 1 | 1 | 1 | 1,88 | 28 |
| LHON | 1 | 1 | 1 | 0,61 | 23 |
| Trial 2 | AUC | Ss | Sp | ES | N |
| SD | 0,91 | 0,79 | 0,93 | 0,04 | 28 |
| RP | 1 | 1 | 1 | 1,75 | 28 |
| LHON | 1 | 1 | 1 | 0,72 | 23 |

**Figure 2:**
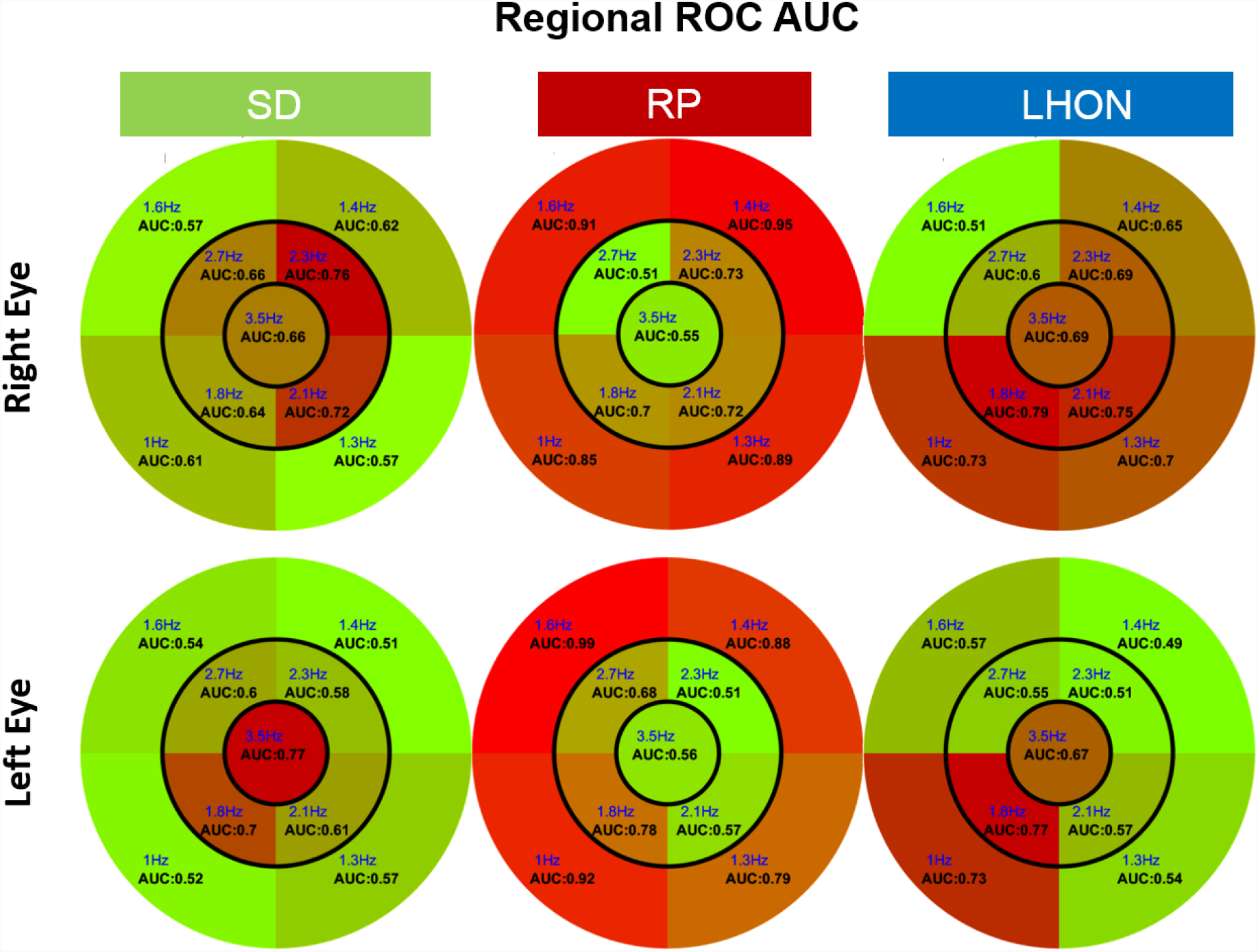
AUC of ROC for each sector of the large field stimulus: AUC, Sensitivity (Ss), specificity (Sp) for SD, RP and LHON patients relative to HP participants. AUC of ROC are computed for each TMF/stimulus sector and each pathology. The distribution of AUC values, indicated in black on each stimulus sector, maps the regional visual defects identified with MFPT. The numbers in blue indicate the tagging frequency of each sector.

To determine whether SPPf distribution differentiates diseases from one another, we further computed ROC curves contrasting RP from SD patients, RP from LHON patients and SD from LHON patients. AUC of ROC curves computed with the spectral power of all FOIs was greater than 0.90 for all comparisons (Supplementary table 3).

The above results were obtained by pooling data from two trials separated by a delay of 45 to 60 minutes, during which several other pupillary tests were performed. Sensitivity and specificity computed separately for each trial were similarly high (Table 1). Test-retest stability was computed as Pearson’s correlation coefficients between trials and Bland-Altman plots^24^. Despite fatigue related to the lengthy session of our protocol comprising many different stimuli (about 2 hours), MPFT test-retest repeatability was good in healthy participants (r=0.81, p<0.001), SD (SD: r=0.8, p<0.001) and LHON (r=0.77, p<0.001), but lower in the RP patients (RP: r=0.69, p<0.001; Supplementary Figure 2).

We then determined whether visual defects could be localized with greater spatial resolution with MFPT. To that aim, we designed two multipartite stimuli with 9 small sub-regions covering opposite visual quadrants (see Methods and Supplementary Figure 3), presented in two successive 45 second trials. Again, the SPPf allowed sorting patients from healthy participants with high sensitivity and specificity at both global and local scales (Supplementary Figure 3B), demonstrating that a coarse-to-fine strategy allows localizing visual defects at high resolution, if the large-field stimulation revealed abnormal pupillary responses.

As it can be seen in Table 1, AUC, sensitivity and specificity were somewhat lower for patients with LHON and SD than for RP patients. This possibly reflects a greater heterogeneity of visual defects across participants, and the fact that LHON and SD patients with central visual defects are more prone to drifting their fixation and making saccades. Furthermore, a detailed analysis of the raw pupillary responses revealed that the pupil of healthy participants slowly dilates over the course of a run, reminiscent of the pupillary “escape” seen during long lasting stimulation^25^. This trend was less pronounced in RP, SD and LHON patients (Supplementary Figure). Consequently, including the slopes of this linear trend in the ROC analyzes improved the classification of patients (Supplementary Table 3).

To determine whether the functional pupillary responses relate to the structural deficits, we computed the correlation coefficients between the SPPf and retinal thickness (RNFL) measured by optical coherence tomography (OCT, see Methods), at 3 different eccentricities. The two variables were correlated significantly (r=0.73, p<0.001) in healthy participants, suggesting that pupillary responses can provide a proxy to retinal thickness. The correlation coefficients were not as high in participants with retinal disease (RP, r=0.45, p<0.012; SD, r=0.56, p<0.02) and not significant in optic nerve disease (LHON, r=0.42, p<0.11; Supplementary Figure 4), possibly because retinal thickness as measured herein does not distinguish between different retinal layers (although this information existed in the data set, it was not possible to retrieve it in the present study).

## Discussion

Overall, the present MFPT proof-of-concept shows that sustained pupillary responses to a multipartite stimulus with frequency-tagged luminance modulations has the potential to quickly and reliably assess the existence of localized visual field defects. Probing functional defects with MFPT is advantageous because it is very fast and tests the visual field at once. This simultaneous pupillary assessment further permits evaluating the *relative* spectral power between different FOIs at once, rather than during successive stimulations. This is important because comparing pupillary responses elicited in succession over a long duration may be biased by changes in pupillary responsivity over time. In addition, the standard deviations of the relative spectral power across different regions provides an additional measure for characterizing diseases, as expected when specific regions are impaired (see Supplementary Figure 5). Finally, the *relative* evaluation of pupillary responses can be evaluated independently of systemic factors such as fatigue, stress, attention, drugs or medication that may modulate pupillary responses, avoiding burdening participants to control for these potential confounds. As MFPT elicits sustained pupillary responses, it is robust to blinks or saccades artifacts that may transiently perturb pupillary activity, as long as these artifacts are not too frequent and their overall duration is not too long (note that no trial had to be removed from the analyzes because of spurious data).

Most often, pupillographic perimetry uses numerous brief bright light stimulations in succession, to elicit series of Pupil Light Reflex (PLR) and/or post illumination pupil responses (PIPR). One study reports using frequency-tagging, but with a single frequency^26^.These methods often require longer examination time than MFPT, can dazzle the participants with sudden bright flashes, and may be more prone to contamination by eye-movements, blinks, fatigue and cognitive factors. As a matter of fact, the discrete activity elicited with many brief light flashes followed by periods (PIPR) during which participants are presented a dark background may foster mind wandering or attention lapses. With MFPT, pupillary activity is continuously constrained by luminance oscillations, such that all pupillary and eye-movement data are relevant to assess the existence of visual defects. In this regard, the finding of a slow pupillary dilation over the course of a trial brings additional evidence to distinguish impaired patients from healthy participants. Although attentional lapses or focused attention onto a sector could modulate pupillary activity^27^, these pupillary modulations are unlikely to counterbalance the continuous strong visual drive of MFPT. For attention to significantly modulate pupillary activity would require sustained attention –to one or several sectors-during the whole test, which appears unlikely. To nevertheless test for this possibility, we conducted experiments with young healthy participants, whose task was to report whether two colored disks displayed side-by-side onto one sector had the same hue. The colors of the disks changed every second. Each of the 45 seconds trials (one for each sector) thus required attending to a single location during the whole duration of the test. A control condition used the same stimuli, but with no associated task (passive fixation). Although we did found a significant increase of the mean spectral power in the attention, relative to the no task, condition (p<0.02), we did not found significant selective increases of the spectral power associated to the sector where the colored disk stimuli were displayed (unpublished report, 2018).

In contrast to threshold Standard Automatised Perimetry that relies on subjective criteria-dependent responses, MFPT is an objective physiological measurement of reflexive sustained pupillary activity that can be administered without particular expertise, opening the way to routinely screen and follow-up visual defects in a variety of ophthalmologic diseases, whose prevalence worryingly increases worldwide. Although the spatial resolution of MFPT is coarser than SAP, it is well suited to an adaptive coarse-to-fine strategy, allowing identifying visual defects at progressively finer spatial scales, when needed. Importantly, MFPT is nearly effortless for subjects, requires no volitional responses, and does not dazzle the participants at it uses slow luminance modulations of moderate amplitude. Thus, MFPT is suitable for all populations –including young children, elderly with cognitive deficits, non-human mammals-having difficulties understanding instructions, performing subjective tasks, or sustaining fixation and attention over extended periods of time.

This proof-of-concept study has limitations: first, the small number of participants and the choice or rare diseases affecting mostly young adults precludes any generalization to other diseases and older individuals. The severity of the diseases of the patients included in the study was heterogeneous such that, given the small number of participants, it was not possible to analyze pupillary responses as a function of the state of the disease. Second, the displays used herein are grey, and therefore stimulate all classes of photoreceptors as well as intrinsically responsive ganglion cells (ipRGCS) containing melanopsin sensitive to blue light. Therefore we could not separately probe the different retinal circuits –direct ipRGCs stimulation versus indirect stimulation through cones/rods circuits that are involved in pupillary activity. Third, the size of the display (∼40°) was limited such that the far periphery could not be tested. Finally, in this study, we did not correct for drifting fixation that may have occurred and bias the results.

The present proof-of-concept study conducted on few patients and rare diseases suggests MFPT has the potential to quickly assess visual defects, with little burden for the patients. Importantly, for the patients with an optic neuropathy, the mean spectral power of pupillary responses is correlated to the structural damage (RNFL) measured with OCT, in agreement with other studies^12^. Whether this correlation exists at or before disease onset remains to be investigated in longitudinal studies and more frequent pathologies (Glaucoma, AMD or diabetic retinopathy).

In this regard, an ongoing study with Glaucoma patients suggests that the sensitivity, specificity and reliability of MPFT are similar to those reported herein, and that MPFT is better accepted than SAP. These preliminary results also indicate that the pupillary spectral power is correlated to RNFL. To conclude, MFPT could be used for screening populations at risk, or to follow-up the evolution of ophthalmologic diseases, as well as neurologic pathologies where pupillary responses were found to be altered^28,29^. However, screening or following-up evolving diseases require more clinical evidence on larger populations and more frequent diseases to determine a discriminant pupillary threshold that reliably distinguishes patients from healthy individuals.

## Material and Method

### Participants

Fourteen patients with Retinitis Pigmentosa (RP: mean age: 41, sd: 11; 6 women), 14 patients with Stargardt disease (SD: mean age: 38, sd: 9; 5 women), 9 patients with Leber hereditary optic neuropathy (LHON: mean age: 33, sd: 7; 4 women) and 14 healthy participants (HP: mean age: 37, sd: 10; 6 women) were included in the study. All participants were aged between 20 and 58 years. The inclusion criteria for the healthy volunteers were a binocular corrected visual acuity greater or equal to 8/10 (≤ 0.1 logMAR) and a normal visual field (Supplementary table 1). Patients and healthy participants were tested without corrections (lens or glasses) during the main experiment.

### Protocol

All participants passed a full ophthalmic assessment, including visual acuity (ETDRS right and left eye, and binocular), color vision test (saturated and desaturated 15 Hue tests: D-15d), visual field test (Humphrey Field Analyzers, HFA, model Octopus 900, using isopter V4 to III), contrast sensitivity (Pelli-Robson, left, right eye and binocular), fundus examination as well as a macular (macular volume and thickness) and optic nerve (peripapillary retinal nerve fiber layer thickness) optical coherence tomography (OCT, Spectralis Heidelberg, Engineering, Heidelberg, Germany). Whenever possible, the ophthalmic assessment was performed on the same day as the pupillary tests.

### Apparatus

The stimuli were back-projected at 60 Hz using a Titan 1080p video projector (Digital Projection, Ldt) on a translucent screen (55,5 × 41.63 degrees of visual angle). Participants were conformably installed at 128 cm from the screen, while resting their head on a chin-rest. Pupil modulations and eye-movements of both eyes were recorded with an eye-tracker (EyeLink II, 500 frames/s., SR Research, Ldt), and down-sampled to 60 frames/s. Each eye could be stimulated independently, using a large mobile black cardboard that masked the display screen to one eye. Dedicated custom software (Jeda) was used for stimulus generation and eye-movement recordings. (Note that similar results were found in other studies using a conventional monitor and a different eye-tracker).

### Stimuli

Three different spatial distributions of annulus-sectors were used, with each sector being coupled with a sinusoidal luminance modulation at a specific temporal frequency (TMF). One, large-field, stimulus was composed of 9 sectors (Figure 1A) whose locations, separations and relative sizes, as well as the values and distribution of the TMFs and stimulation duration, were chosen based on in-depth preliminary studies conducted on healthy participants. The choice of the sinusoidal TMF was dictated by several considerations: 1. the highest TMF in the stimulus was lower than 4 Hz to ensure that reliable and sustained pupil responses would be present. 2. The lowest TMF was around 1 Hz, to ensure that a too limited number of cycles during a trial would not bias the spectral analyzes. 3. TMFs were chosen to be incommensurate, so as to avoid that harmonics and fundamental frequencies overlap, as this could introduce artifacts and alter the analyses of the pupil responses. In addition, care was taken to avoid FOIs corresponding to intermodulation products, corresponding to the sum and difference between frequencies that may appear in the Fourier spectrum, if non-linear interactions between frequencies would occur. 4. Finally, the selection of TMF was constrained by the screen refresh rate, (which was kept to 60 Hz, to be compatible with any monitor). The 4 lowest TMFs were associated to the 4 eccentric annulus sectors, the 4 intermediate TMFs were associated to the para-central annulus sectors. The highest TMF was presented in central vision to weight the, otherwise dominant, contribution of central vision to pupillary activity^30,31^, which also helped maintaining fixation over time. As a result of these constraints, the distribution of TMFs was: 1.00 1.25 1.39 1.58 1.81 2.14 2.31 2.73 and 3.56 Hz, with the periods of each TMF corresponding to an integer number of frames at 60 Hz: 60, 48, 43, 38, 33, 28, 26, 22, 17 frames. The minimum duration of the sustained stimulation that still evoked reliable pupillary responses at all FOIs (45 sec.) was assessed in preliminary experiments (Note that longer durations increase the frequency resolution and enhance the power spectrum at FOIs).

In addition to this large-field stimulus, we designed two stimuli with 9 annulus sectors covering 2 opposite quadrants of the visual fields (Left-up + Right-down and Right-up + Left-down; see supplementary Figure 3A). The TMFs of these Quadrant stimuli were identical to those of the large-field stimulus.

The amplitude of luminance modulations, chosen on the basis of preliminary experiments so as to avoid glare while still providing reliable power in the Fourier spectrum, were identical for all sectors and stimuli and ranged from 65 cd/m^2^ to 181.10 cd/m^2^, with an averaged mean luminance of 123 cd/m^2^. (Note that using the same design with graphics displays having different luminance settings gave results similar to those described herein).

The large-field stimulus covered 40.6 degrees of visual angle (central region: 4.6°, paracentral sub-regions 19.6°). Central, paracentral and eccentric sectors were separated by black stripes (width 0.5°). The quadrant stimuli covered the same spatial extent, except that only 2 opposite quadrants (each made of 4 sectors) were visible during a trial. Sectors were separated by dark, 0.3° wide gaps. We used 2 opposite quadrants to limit involuntary eye-movements or biased fixation that may occur with a single quadrant.

### Procedure

The session started with positioning the participants and adjusting the cameras of the eye-tracker. After 10 minutes of adaptation to the low ambient light of the testing room, during which a questionnaire was given to the participants, a 5 point eye calibration procedure was given before series of short (<2 minutes) tests, including PLR measures, RAPD assessment, as well as other tests, including the large-field and quadrant MFPT tests. For all these tests, participants were simply asked to maintain fixation at the center of the screen, marked by a small white circular fixation point, with no other concurrent task. A brief rest, adapted to each participant, separated the different runs, and was used when necessary to change the stimulated eye (Right or Left; note that eye-movements and pupillary responses of both eyes were always recorded during a run). To evaluate test-retest variability, the large-field and quadrant MFPT tests were repeated twice for each eye, with a delay of about 45 to 60 minutes between recordings, during which other pupillary tests were given to the participants. Overall, a session lasted about 2 hours, but the MFPT tests (2 eyes x 2 repetitions for the large field MFPT; 2 eyes x 2 trials for the quadrant MFPT) only took about 12 min. A MFPT test started with a brief full screen flash (150 msec.) at the maximum screen luminance, (181.10 cd/m^2^) followed by a dark screen for 3 seconds, before the static MFPT grey stimulus appeared on the screen for 3 seconds, followed by 45 second of temporal modulations. In this way, a PLR was recorded as a reference baseline before each MFPT run.

### Data analyzes

All recorded data were included in analyzes, performed with Matlab R2018a (The MathWorks, Natick, MA, USA). Data for the second trial of one SD patient were lost due to technical issues. The raw pupil data were down-sampled at 60 Hz and corrected for blinks and artifacts by replacing lacking or spurious data by a smoothed linear interpolation, using a pre- and post-blink offset of 4 samples (66.6 msec). Blinks and spurious data were detected using both a velocity and an acceleration thresholds, chosen once for the whole data set. As the initial and final pupillary responses were often noisy, the signal was trimmed by 120 samples (3 sec.), so that 39 seconds of pupillary responses were used for the analyses. The corrected pupillary signals were then z-scored, and a Fast Fourier transform (FFT) was computed on the so-corrected pupillary signals. The resulting power spectrum was normalized by a “regional” mean power spectrum for each eccentricity of the multipartite stimulus. This was done by dividing the power spectrum for the low TMF/Peripheral sectors (1, 1.25, 1.39 and 1.58 Hz) by the mean power spectrum between 0.88 and 1.74 Hz; the medium TMF/Paracentral sectors (1.81, 2.14, 2.3 and 2.72 Hz) by the mean power spectrum between 1.76 and 3 Hz; and the high TMF/Central sector (3.5 Hz) by the mean power spectrum between (3.02 and 3.94 Hz). Normalizing the power spectra in this way takes into account the spurious noisy power that can exist for each frequency band (Supplementary Figure 7A).

We ascertain that peaks in individual spectral power were triggered by, and phase locked to, the stimulus FOIs, by comparing the spectral power of the average pupillary responses to the average of individual power spectra. If individual pupillary responses were not phase locked to FOIs, phase shifts would flattened the mean pupil response, whose power spectrum would lack peaks at FOIs. This was not the case (Supplementary Figure 7B), demonstrating that stimulus FOIs did tag the pupillary response in each case, although with varying power depending on the disease at stake. Time Frequency maps were also computed to verify that, on average, the power spectrum at FOIs was sustained during a run and did not result from short episodes of oscillatory activity (data not shown; see examples in supplementary Figure 8).

We used the spectral FOI power distribution to compute the area under the receiving operating characteristic curves (AUC of ROC). The area under the curve (AUC) and the corresponding sensitivity, Ss, and specificity, Sp, were computed using the *fitglm* and *perfcurve* Matlab functions: first, a logistic regression model was first computed with the Matlab *fitglm* function using the power spectrum at all FOIs. Second, the probability distribution returned by the *fitglm* function as scores was then used to compute the ROC curves with the Matlab function *perfcurve*.

ROC curves were computed separately for each eye using: **i**. the spectral power of all FOIs from both trials to discriminate the different patient groups (SD, RP and LHON) from healthy participants (HP), **ii**. The spectral power of all FOIs of each eye and each trial **iii**. AUC of ROC were also computed with the spectral power of each eye at each frequency-tagged sub-region of the large-field and quadrant multipartite stimuli, resulting in a synthetic pupillary field maps describing the power distribution over space for each participant, as well as the corresponding AUC distribution of ROC for group analyses (Figure 2).

Test-retest variability was assessed by computing the Pearson’s coefficient correlation between the normalized FOI spectral power collected during the first and second MFPT trials (ran with a delay of about >60 min), as well as Bland-Altman plots.

To compute the correlations between the retinal thickness measured with OCT and the spectral power, we matched as much as possible the eccentricities of both measures and used the averaged values at each eccentricity. Note, however, that OCT measures and MFPT measures could not perfectly superimpose (See Supplementary Figure 1).

## Data Availability

All data produced in the present study are available upon reasonable request to the authors

## Acknowledgments

The authors are very grateful to all the participants who accepted to contribute to this study, and to Patrick Cavanagh, Mark Wexler and Jochen Braun for their suggestions and comments on an earlier version of the manuscript. The study was funded by the “Fondation Voir & Entendre”, and took place in the “Street Lab” facilities at “Institut de la Vision”.

## Author contributions

JL and CV wrote the manuscript; JL wrote software for visual stimulation and data acquisition; SA, CC and JL defined the protocol; SA and JL analyzed the data; CV and CC conducted the patients’ examination; MS and CR managed the recruitment of the patients.

## Ethics declarations

All methods were carried out in accordance with relevant guidelines and regulations. All experimental protocols were approved by a named institutional and/or licensing committee/s. The protocol conformed to the Declaration of Helsinki and was approved by the French local ethic committee (“Comité de Protection des Personnes Ile de France XI”, Ref CPP: 18002, Code: P1705, ID-RCB: 2017-A03236-47; NCT04909398). All participants gave their written informed consent to participate in the study, and were free to leave the study whenever they decided to do so.

## References

1. Wong, W. L. et al. Global prevalence of age-related macular degeneration and disease burden projection for 2020 and 2040: a systematic review and meta-analysis. Lancet Glob. Health 2, e106–e116 (2014).

2. Tham, Y.-C. et al. Global Prevalence of Glaucoma and Projections of Glaucoma Burden through 2040. Ophthalmology 121, 2081–2090 (2014).

3. Kerrigan, L. A., Quigley, H. A., Pease, M. E., Kerrigan, D. F. & Mitchell, R. S. Number of Ganglion Cells in Glaucoma Eyes Compared with Threshold Visual Field Tests in the Same Persons. 41, 8 (2000).

4. Gardiner, S. K., Mansberger, S. L. & Fortune, B. Time Lag Between Functional Change and Loss of Retinal Nerve Fiber Layer in Glaucoma. Investig. Opthalmology Vis. Sci. 61, 5 (2020).

5. Gardiner, S. K. & Demirel, S. Assessment of Patient Opinions of Different Clinical Tests Used in the Management of Glaucoma. Ophthalmology 115, 2127–2131 (2008).

6. Werner, E. D. Variability of Automated Visual Fields in Clinically Stable Glaucoma Patients. 30, 7.

7. Kucur, S. S. et al. Comparative Study Between the SORS and Dynamic Strategy Visual Field Testing Methods on Glaucomatous and Healthy Subjects. Transl. Vis. Sci. Technol. 9, 3 (2020).

8. Bergamin, O. & Kardon, R. H. Latency of the Pupil Light Reflex: Sample Rate, Stimulus Intensity, and Variation in Normal Subjects. Investig. Opthalmology Vis. Sci. 44, 1546 (2003).

9. Maddess, T., Bedford, S. M., Goh, X.-L. & James, A. C. Multifocal pupillographic visual field testing in glaucoma. Clin. Experiment. Ophthalmol. 37, 678–686 (2009).

10. Carle, C. F., James, A. C., Kolic, M., Essex, R. W. & Maddess, T. Blue Multifocal Pupillographic Objective Perimetry in Glaucoma. Investig. Opthalmology Vis. Sci. 56, 6394 (2015).

11. Sabeti, F., James, A. C., Essex, R. W. & Maddess, T. Multifocal pupillography identifies retinal dysfunction in early age-related macular degeneration. Graefes Arch. Clin. Exp. Ophthalmol. 251, 1707–1716 (2013).

12. Najjar, R. P. et al. Pupillary Responses to Full-Field Chromatic Stimuli Are Reduced in Patients with Early-Stage Primary Open-Angle Glaucoma. Ophthalmology 125, 1362–1371 (2018).

13. Adhikari, P., Zele, A. J., Thomas, R. & Feigl, B. Quadrant Field Pupillometry Detects Melanopsin Dysfunction in Glaucoma Suspects and Early Glaucoma. Sci. Rep. 6, 33373 (2016).

14. Jain, M. et al. Pupillary Abnormalities with Varying Severity of Diabetic Retinopathy. Sci. Rep. 8, 5636 (2018).

15. Barbur, J. L., Moro, S., Harlow, J. A., Lam, B. L. & Liu, M. Comparison of pupil responses to luminance and colour in severe optic neuritis. Clin. Neurophysiol. 115, 2650–2658 (2004).

16. Rao, H. L. et al. Predicting the intereye asymmetry in functional and structural damage in glaucoma using automated pupillography. Acta Ophthalmol. (Copenh.) 95, e532–e538 (2017).

17. Chang, D. S. et al. Symmetry of the Pupillary Light Reflex and Its Relationship to Retinal Nerve Fiber Layer Thickness and Visual Field Defect. Investig. Opthalmology Vis. Sci. 54, 5596 (2013).

18. Erdem, S. et al. The effectiveness of automatic pupillometry as a screening method to detect diabetic autonomic neuropathy. Int. Ophthalmol. 40, 3127–3134 (2020).

19. Sabeti, F. et al. Comparing multifocal pupillographic objective perimetry (mfPOP) and multifocal visual evoked potentials (mfVEP) in retinal diseases. Sci. Rep. 7, 45847 (2017).

20. Sabeti, F., James, A. C. & Maddess, T. Spatial and temporal stimulus variants for multifocal pupillography of the central visual field. Vision Res. 51, 303–310 (2011).

21. Sabeti, F. et al. Objective testing of both visual fields in 80 seconds. 1.

22. Ajasse, S. Pupillométrie dynamique : approches fondamentales et cliniques. Sciences cognitives. (Sorbonne Université, 2019).

23. Schmid, R., Wilhelm, B. & Wilhelm, H. Naso-temporal asymmetry and contraction anisocoria in the pupillomotor system. Graefes Arch. Clin. Exp. Ophthalmol. 238, 123–128 (2000).

24. Altman, D. G. & Bland, J. M. Measurement in Medicine: The Analysis of Method Comparison Studies. The Statistician 32, 307 (1983).

25. Adhikari, P., Zele, A. J. & Feigl, B. The Post-Illumination Pupil Response (PIPR). Investig. Opthalmology Vis. Sci. 56, 3838 (2015).

26. Naber, M. et al. Gaze-Contingent Flicker Pupil Perimetry Detects Scotomas in Patients With Cerebral Visual Impairments or Glaucoma. Front. Neurol. 9, 558 (2018).

27. Rosli, Y. et al. Retinotopic effects of visual attention revealed by dichoptic multifocal pupillography. Sci. Rep. 8, 2991 (2018).

28. Tabashum, T. et al. Detection of Parkinson’s Disease Through Automated Pupil Tracking of the Post-illumination Pupillary Response. Front. Med. 8, 645293 (2021).

29. Nakayama, M., Nowak, W., Krecicki, T. & Hachol, A. Prediction of Alzheimer’s Disease in Patients using Features of Pupil Light Reflex to Chromatic Stimuli. in 273–277 (2018). doi:10.15439/2018F57.

30. Hong, S., Narkiewicz, J. & Kardon, R. H. Comparison of Pupil Perimetry and Visual Perimetry in Normal Eyes: Decibel Sensitivity and Variability. 42, 9 (2001).

31. Tan, L., Kondo, M., Sato, M., Kondo, N. & Miyake, Y. Multifocal pupillary light response fields in normal subjects and patients with visual field defects. Vision Res. 41, 1073–1084 (2001).

