## Supplementary Figures and Tables for "One minute Multiple Pupillary Frequency Tagging test to assess visual field defects"

### Supplementary Table 1

| General | | | Refraction RE | | | Refraction LE | | | Visual Acuity (LogMAR) | | | Pelli-Robson (logCS) | | | Summary | |
|---|---|---|---|---|---|---|---|---|---|---|---|---|---|---|---|---|
| n° | Sex | Pathology | Sphere | CYL | Axis | Sphere | CYL | Axis | Right Eye | Left Eye | Binocular | Right Eye | Left Eye | Binocular | Age | Sex |
| 22 | M | HP | NA | NA | NA | NA | NA | NA | -0,3 | -0,2 | -0,3 | 1,65 | 1,65 | 1,65 | 37 +-10 | 8M / 6F |
| 25 | M | HP | NA | NA | NA | NA | NA | NA | -0,2 | -0,2 | -0,24 | 1,95 | 1,95 | 1,95 | Max 58 | |
| 29 | M | HP | -2,5 | 0 | 0 | -2,25 | -0,5 | 125 | 0 | -0,08 | -0,1 | 1,95 | 1,95 | 1,95 | Min 24 | |
| 30 | M | HP | 0 | 0 | 0 | 0 | 0 | 0 | -0,1 | -0,1 | -0,2 | 1,95 | 1,95 | 1,95 | | |
| 35 | F | HP | 0 | 0 | 0 | 0 | 0 | 0 | -0,02 | -0,1 | -0,12 | 1,95 | 1,95 | 1,95 | | |
| 36 | M | HP | -2 | -0,5 | 84 | -1,5 | -0,5 | 100 | -0,16 | -0,16 | -0,2 | 1,95 | 1,95 | 1,95 | | |
| 37 | F | HP | -10,25 | -1,25 | 157 | -9,5 | -1,75 | 172 | 0 | -0,04 | -0,1 | 1,95 | 1,95 | 1,95 | | |
| 38 | F | HP | 0 | 0 | 0 | 0 | 0 | 0 | -0,16 | -0,12 | -0,18 | 1,95 | 1,95 | 1,95 | | |
| 39 | F | HP | 0 | 0 | 0 | 0 | 0 | 0 | 0 | 0,04 | -0,04 | 1,95 | 1,95 | 1,95 | | |
| 40 | M | HP | 0 | 0 | 0 | 0 | 0 | 0 | -0,06 | -0,1 | -0,2 | 1,95 | 1,95 | 1,95 | | |
| 44 | M | HP | 0 | 0 | 0 | 0 | 0 | 0 | -0,16 | -0,2 | -0,26 | 1,95 | 1,95 | 1,95 | | |
| 46 | F | HP | -5,5 | -0,5 | 166 | -5 | -1,5 | 43 | 0 | -0,1 | -0,1 | 1,95 | 1,95 | 1,95 | | |
| 54 | F | HP | 0,5 | -0,25 | 106 | 0,75 | 0 | 0 | -0,2 | -0,12 | -0,26 | 1,95 | 1,95 | 1,95 | | |
| 55 | M | HP | 0 | 0 | 0 | 0 | 0 | 0 | -0,3 | -0,3 | -0,3 | 1,95 | 1,95 | 1,95 | | |
| 3 | M | SD | +0,5 | 0,25 | 80 | +0,5 | 0,25 | 175 | 0,86 | 0,76 | 0,78 | 1,65 | 1,65 | 1,65 | 38+-9 | 10M / 4F |
| 4 | M | SD | -0,25 | 0,25 | 65 | -0,75 | 0,25 | 5 | 0,3 | 0,4 | 0,2 | 1,5 | 1,35 | 1,65 | Max 55 | |
| 12 | F | SD | -2 | -0,5 | 75 | -2,25 | -1 | 90 | 1,02 | 1,02 | 1 | 1,2 | 1,2 | 1,2 | Min 21 | |
| 15 | F | SD | -1,25 | -0,5 | 80 | -1,75 | -0,75 | 25 | 0,86 | 0,74 | 0,74 | 1,5 | 1,65 | 1,5 | | |
| 27 | M | SD | -1,75 | -0,75 | 20 | -2 | -0,5 | 160 | 0,9 | 0,9 | 0,9 | 1,35 | 1,35 | 1,35 | | |
| 28 | M | SD | 1,75 | -1,5 | 25 | 2,5 | -2,25 | 170 | 1 | 1,1 | 1,1 | 1,35 | 1,35 | 1,35 | | |
| 31 | M | SD | -2,5 | -1,75 | 170 | -1,5 | -2 | 175 | 0,9 | 0,9 | 0,9 | 1,5 | 1,65 | 1,65 | | |
| 32 | M | SD | -1,5 | -1,75 | 180 | -1,25 | -3 | 180 | 0,94 | 0,96 | 0,86 | 1,5 | 1,2 | 1,5 | | |
| 41 | F | SD | 4 | -2 | 180 | 5 | -2,25 | 5 | 1,04 | 1,18 | 1 | 1,5 | 0,9 | 1,35 | | |
| 42 | M | SD | 5,5 | -1,75 | 15 | 7 | -0,5 | 7 | 1,26 | 1,16 | - | 0,75 | 0,6 | DM | | |
| 43 | M | SD | 0 | -1,5 | 170 | -1,5 | -0,5 | 170 | 1,04 | 0,92 | 0,92 | 1,05 | 1,2 | 1,35 | | |
| 50 | M | SD | -3 | -0,75 | 105 | -3,5 | -0,25 | 140 | 1 | 1,16 | 1,02 | 1,35 | 1,2 | 1,35 | | |
| 51 | F | SD | 0,5 | -1,25 | 25 | 0,75 | -1,25 | 150 | 1 | 1 | 1 | 1,05 | 1,05 | 1,2 | | |
| 53 | M | SD | 0,75 | -0,75 | 90 | 0,75 | -0,5 | 115 | 1,1 | 1,3 | 1,2 | 1,2 | 1,35 | 1,35 | | |
| 2 | F | RP | -1,75 | 2 | 5 | -1,75 | 2,5 | 165 | 0,26 | 0,34 | 0,22 | 0,9 | 0,9 | 1,2 | 41+-10 | 8M / 6 F |
| 6 | M | RP | -7,25 | -2,25 | 20 | -8 | -2 | 175 | 0,38 | 0,42 | 0,42 | 1,65 | 1,35 | 1,65 | Max 58 | 11 |
| 7 | M | RP | -4,75 | 3,25 | 130 | -6,25 | 2,5 | 25 | 0,72 | 0,76 | 0,66 | 0,45 | 0,15 | 0,45 | Min 24 | |
| 8 | M | RP | -5,5 | -0,25 | 160 | -4 | -1,25 | 20 | 0,04 | 0,04 | 0,02 | 1,65 | 1,65 | 1,65 | | |
| 9 | M | RP | 0 | 0,5 | 100 | -0,5 | 0,5 | 110 | -0,04 | -0,08 | -0,08 | 1,65 | 1,65 | 1,65 | | |
| 10 | M | RP | -6 | 1,5 | 10 | -3,5 | 0,75 | 180 | 0,84 | 1,02 | 0,78 | 0,45 | 0,3 | 0,45 | | |
| 11 | M | RP | -0,5 | 1 | 125 | -2 | 1,25 | 10 | 0,72 | 0,7 | 0,68 | 0,45 | 0,6 | 0,6 | | |
| 13 | F | RP | -2,75 | -1,5 | 85 | -3,25 | -1,5 | 80 | 0,06 | 0,22 | 0 | 1,65 | 1,65 | 1,65 | | |
| 14 | M | RP | -1,25 | -1,75 | 90 | 0,75 | -0,5 | 115 | 0,28 | -0,04 | -0,04 | 1,65 | 1,65 | 1,65 | | |
| 16 | F | RP | -0,75 | -1,75 | 75 | -0,75 | -2 | 105 | 0,34 | 0,22 | 0,3 | 1,65 | 1,5 | 1,65 | | |
| 17 | F | RP | -6 | -2,5 | 20 | -6,5 | -2,25 | 155 | 0,34 | 0,44 | 0,34 | 1,35 | 1,05 | 1,35 | | |
| 18 | F | RP | -6 | -2,25 | 15 | -6 | -1 | 135 | 0,22 | 0,12 | 0,1 | 1,65 | 1,5 | 1,65 | | |
| 19 | M | RP | -3,5 | -1 | 0 | -2,25 | -1,25 | 5 | 0,36 | 0,12 | 0,1 | 1,5 | 1,65 | 1,65 | | |
| 24 | F | RP | NA | NA | NA | NA | NA | NA | 0,3 | 0,4 | 0,3 | 1,65 | 1,65 | 1,65 | | |
| 1 | M | LHON | -0,5 | 0,75 | 80 | -0,25 | 1,75 | 90 | 0 | 0,1 | 0 | 1,05 | 1,05 | 1,05 | 33 +-7 | 5M / 4F |
| 20 | F | LHON | -1,5 | -0,25 | 15 | -1 | -0,25 | 15 | 0,32 | - | 0,32 | 0,45 | DM | 0,45 | Max 42 | |
| 23 | M | LHON | 1,25 | NA | NA | 1,25 | NA | NA | 1,52 | 1,48 | 1,48 | 0 | 0,45 | 0,45 | Min 20 | |
| 26 | F | LHON | 2,5 | 0 | 0 | 3,25 | -0,5 | 135 | 1,4 | 1,3 | 1,4 | 0,75 | 1,05 | 0,75 | | |
| 45 | F | LHON | -0,75 | -0,25 | 130 | -0,75 | -0,5 | 15 | 1,02 | 1,1 | 0,92 | 1,05 | 1,05 | 1,35 | | |
| 47 | M | LHON | -3 | -1,25 | 5 | -2,75 | -0,25 | 125 | 1,44 | 1,32 | 1,36 | 0,6 | 0,45 | 0,75 | | |
| 48 | M | LHON | 0,25 | 0 | 0 | 0 | 0 | 0 | 1,04 | 0,24 | 0,44 | 0,15 | 0,15 | 0,15 | | |
| 49 | F | LHON | -0,5 | -0,75 | 100 | -1 | -0,5 | 75 | 1,5 | 1,26 | 1,32 | 0,15 | 0,6 | 0,45 | | |
| 52 | M | LHON | 0 | -0,75 | 10 | 0,25 | -1 | 10 | 1 | 0,96 | 0,96 | 1,35 | 1,5 | 1,65 | | |

Summary statistics of the participants
HP : Healthy participants
SD : Stargardt disease
RP: Retinitis Pigmentosa
LHON : Leber Heriditary Optic Neuropathy

| LE versus RE | AUC | Ss | Sp | N |
| --- | --- | --- | --- | --- |
| HP | 0,97 | 0,89 | 0,96 | 56 |
| SD | 0,95 | 0,93 | 0,89 | 56 |
| RP | 0,69 | 0,96 | 0,39 | 56 |
| LHON | 0,97 | 1 | 0,94 | 36 |
| ALL | 0,97 | 0,94 | 0,87 | 204 |
| ALL-RP | 1 | 1 | 1 | 148 |

AUC of ROC, sensitivity and specificity for discriminating the Left from the Right Eye for HP, RP, SD, LHON, for all participants and all participants but RP patients.

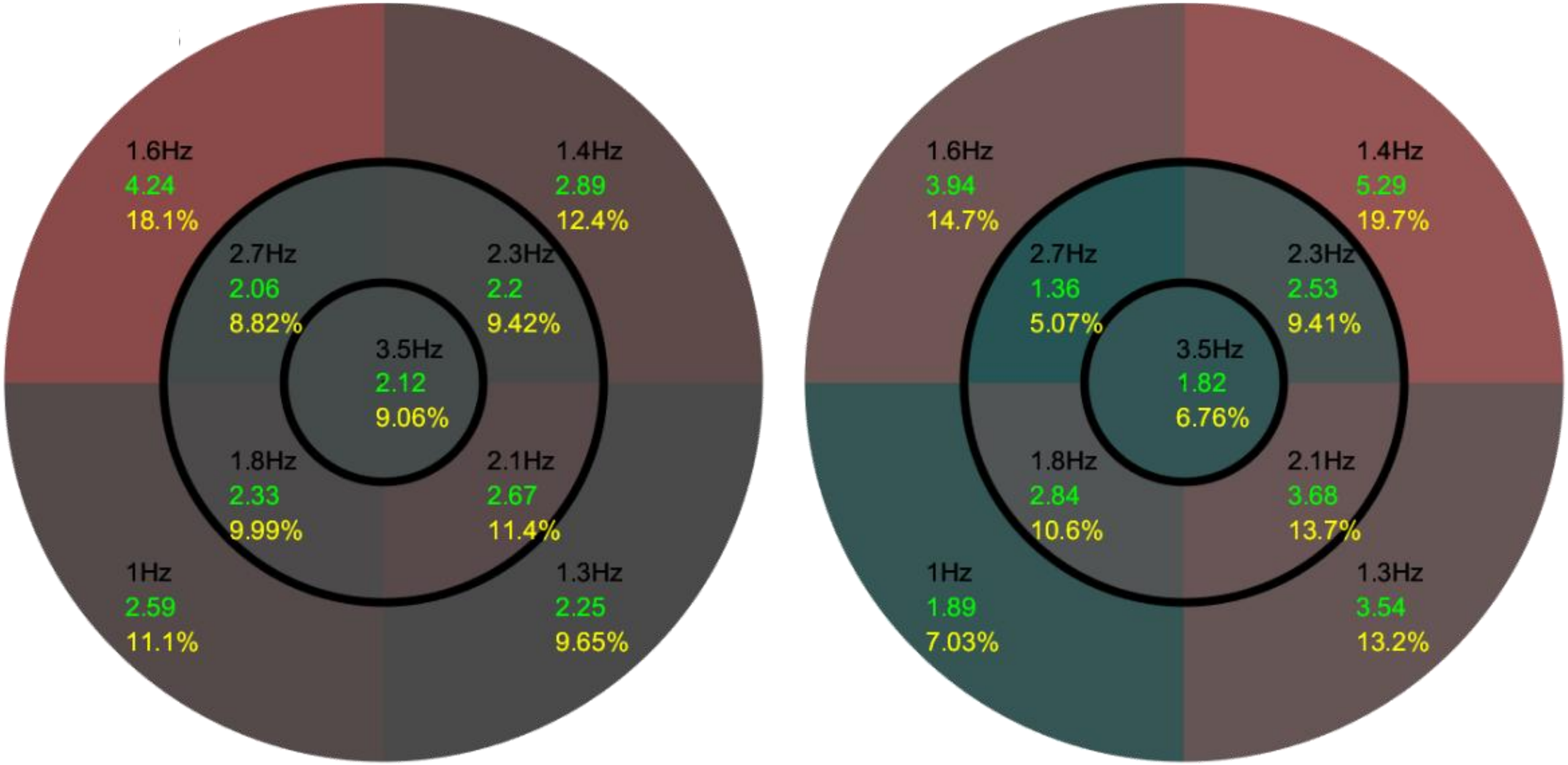


Distribution of the average spectral power across participants for each sector/FOI of the large field stimulus. **Left**: Left Eye; **Right**: Right Eye. *Black values*: temporal modulation frequency for each sector. *Green values* normalized spectral power. *Yellow Values*: respective contribution of each sector to the overall pupillary spectral power, used to set the color code of each sector.

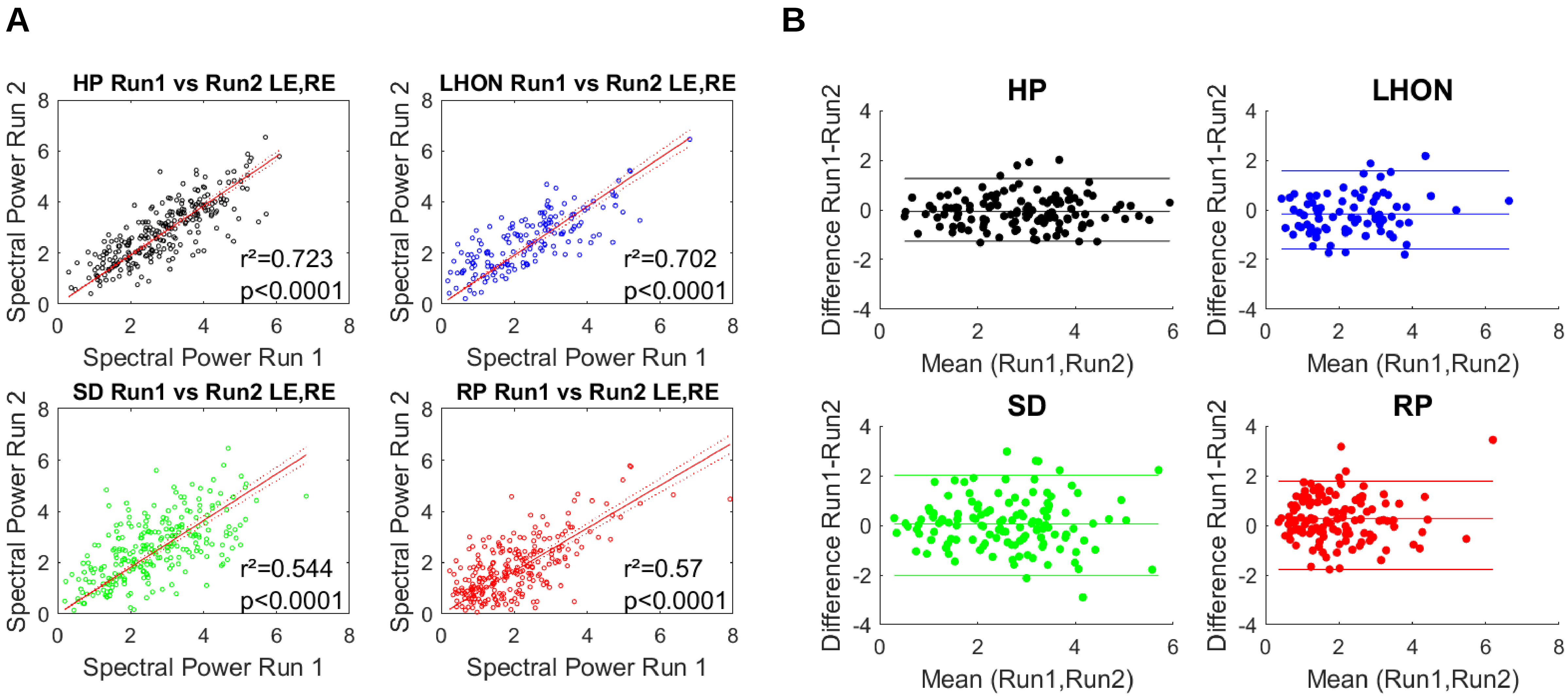


**A**: Test-retest repeatability for each group, evaluated by comparing the spectral power from Run  and Run 2, separated by 45 to 60 min, during which several other pupillary tests we done. **B**: Bland-Altman plots for each group. Horizontal lines indicates the 95% confidence intervals.

**A**

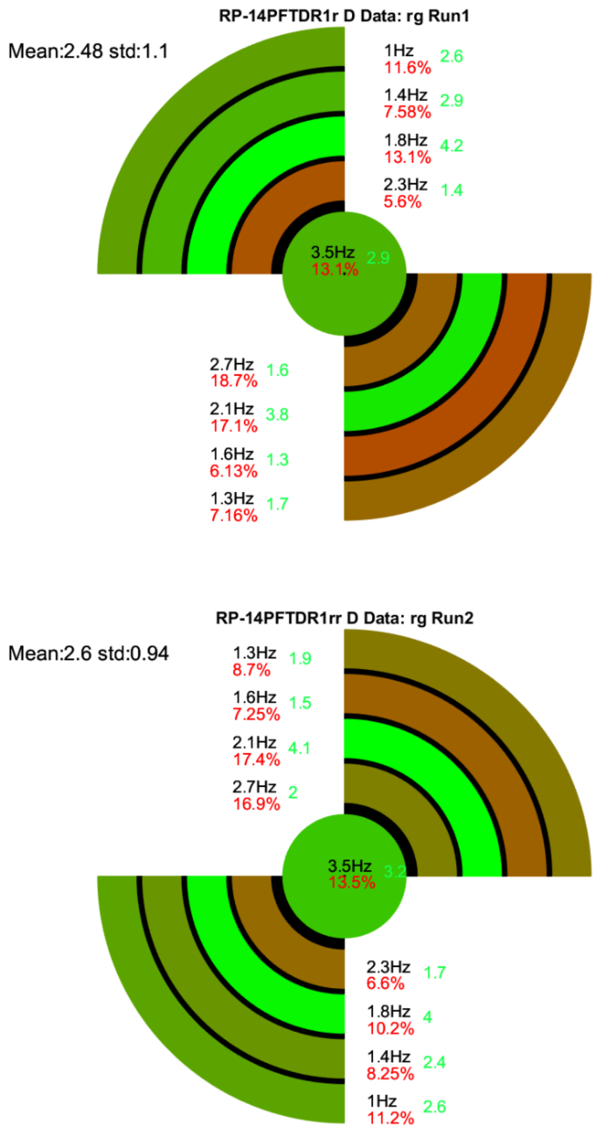


**B**

| RE x 2 Quadrants | AUC | Ss | Sp | N |
|---|---|---|---|---|
| SD | 0,96 | 0,92 | 0,89 | 56 |
| RP | 1 | 1 | 1 | 56 |
| LHON | 0,9 | 0,82 | 0,93 | 46 |
| RE Quadrant 1 | AUC | Ss | Sp | N |
| SD | 1 | 1 | 1 | 28 |
| RP | 1 | 1 | 1 | 28 |
| LHON | 1 | 1 | 1 | 23 |
| RE Quadrant 2 | AUC | Ss | Sp | N |
| SD | 1 | 1 | 1 | 28 |
| RP | 1 | 1 | 1 | 28 |
| LHON | 1 | 1 | 1 | 23 |

| LE x 2 Quadrants | AUC | Ss | Sp | N |
|---|---|---|---|---|
| SD | 0,93 | 0,86 | 0,89 | 56 |
| RP | 1 | 1 | 1 | 54 |
| LHON | 0,9 | 0,88 | 0,86 | 46 |
| LE Quadrant 1 | AUC | Ss | Sp | N |
| SD | 1 | 1 | 1 | 28 |
| RP | 1 | 1 | 1 | 28 |
| LHON | 1 | 1 | 1 | 23 |
| LE Quadrant 2 | AUC | Ss | Sp | N |
| SD | 1 | 1 | 1 | 28 |
| RP | 1 | 1 | 1 | 28 |
| LHON | 1 | 1 | 1 | 23 |

**Results for the Quadrant field stimuli. A**: Illustration of the 2 Quadrant field stimulations tested in 2 separated trials and example of results for a RP patient. *Black values*: Temporal modulation Frequency; *Red values*: relative spectral power; *Green values*: Spectral power for each sector. **B**: AUC of ROC, sensitivity & specificity for the right and left eyes. Results for both quadrant field stimuli and for each quadrant field stimulus.

### Between patients classification scores for the large-field stimulus

| Right Eye x 2 Trials | AUC | Ss | Sp | ES | N |
|---|---|---|---|---|---|
| SD vs. RP | 1 | 1 | 1 | 0,94 | 56 |
| LHON vs. RP | 1 | 1 | 1 | 0,29 | 56 |
| LHON vs. SD | 0,97 | 0,89 | 0,94 | 0,53 | 46 |
| **Left Eye x 2 Trials** | **AUC** | **Ss** | **Sp** | **ES** | **N** |
| SD vs. RP | 1 | 1 | 1 | 2,33 | 56 |
| LHON vs. RP | 1 | 1 | 1 | 1,19 | 56 |
| LHON vs. SD | 0,9 | 0,96 | 0,66 | 1,02 | 46 |

AUC of ROC, Sensitivity, Specificity, Effect Size (Hedges m) differenciating the different pathologies (RP, SD & LHON) computed for each eye using the spectral power of the 9 FOIs of 2 trials. **Top:** Right Eye; **Bottom**: Left Eye. The number of observations is indicated in the last column.

Supplementary Figure 4

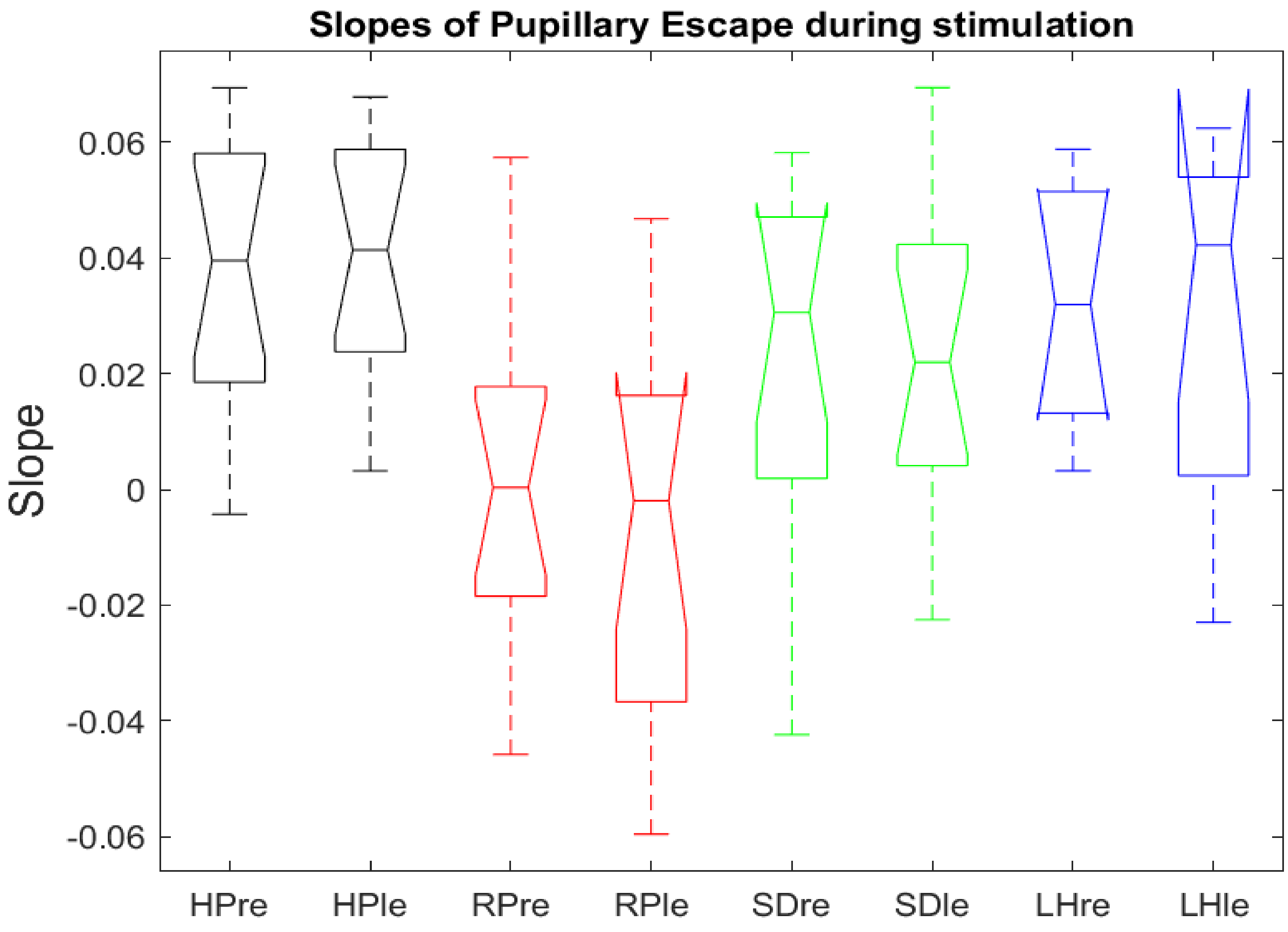


Distribution of the slopes evaluating the pupillary “escape” observed during the visual stimulation for the right and left eyes. Black: HP; Red: RP; Green: SD; Blue, LHON.

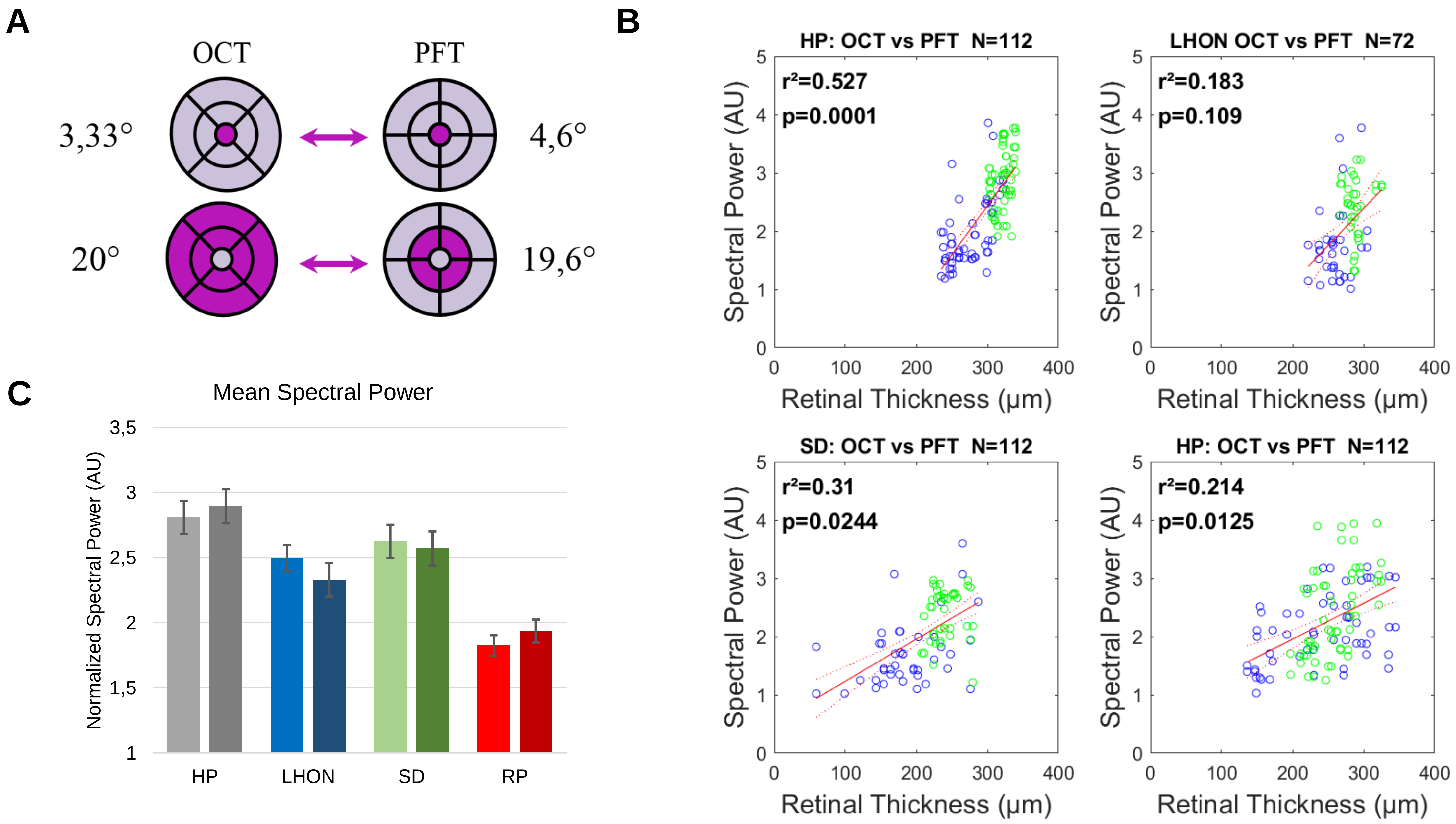


Correlation between Spectral pupillary power and RNFL measured with OCT. **A**. Illustration of the matching between retinal thickness measures and the MPFT stimulus. **B.** Correlations between RNFL and Spectral power for the 4 groups. Green: central measures; Blue: peripheral measures. **C.** Mean spectral power for the right and left eyes of the different groups.

Supplementary Figure 6

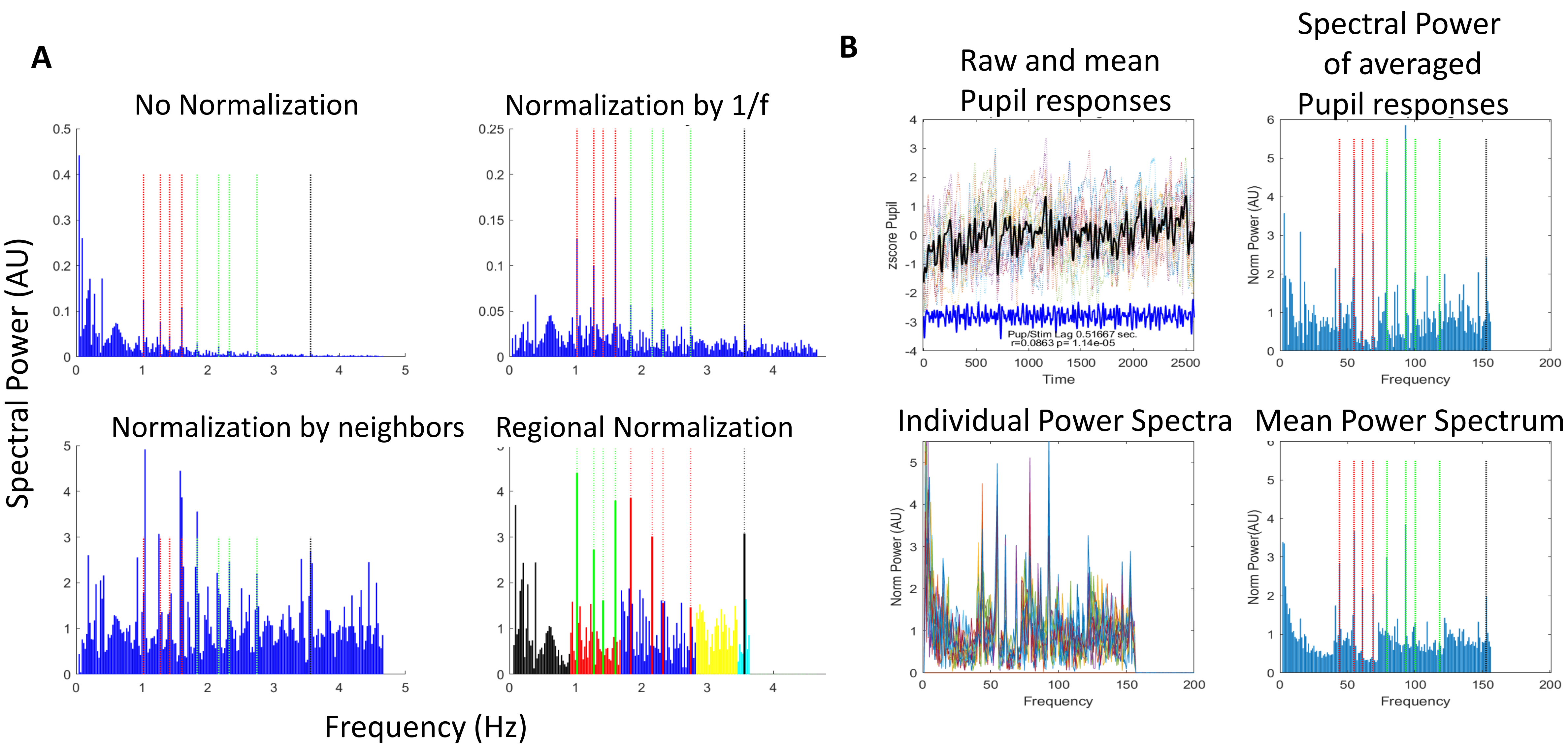


**A.** Different Normalizations of the FFT power. The regional normalization (bottom/right) was used to compute ROC curves (see method for details). **B. Top/Left:** Average pupillary responses across all participants. **Top/Right** : FFT of average pupillary responses. **Bottom/Left:** Spectral power of all particpants. **Bottom/Right:** Average Spectral power across participants. See text for details.

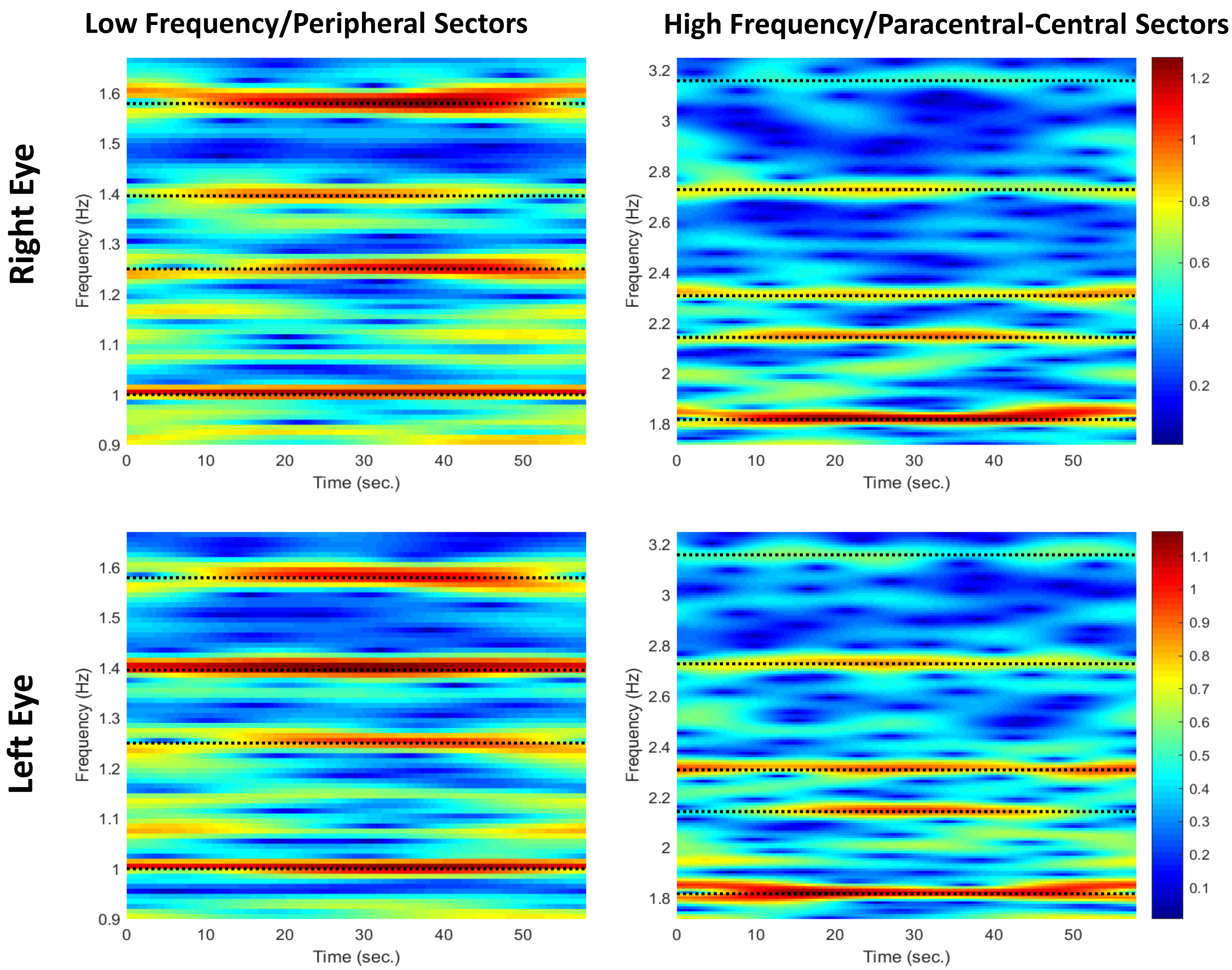


Example of Temporal Frequency Maps of a healthy participant showing sustained pupillary activity at FOIs during the whole stimulation

### Classification scores for the large-field stimulus

| LE+RE x 2 Trials | AUC | Ss | Sp | ES | N |
|---|---|---|---|---|---|
| SD | 0,89 | 0,73 | 0,87 | 0,01 | 111 |
| RP | 0,99 | 0,96 | 0,98 | 1,66 | 112 |
| LHON | 0,86 | 0,67 | 0,93 | 0,74 | 92 |

| LE+RE x 2 Trials + Slopes | AUC | Ss | Sp | ES | N |
|---|---|---|---|---|---|
| SD | 0,93 | 0,83 | 0,88 | 0,01 | 111 |
| RP | 0,99 | 1 | 0,98 | 1,66 | 112 |
| LHON | 0,89 | 0,8 | 0,88 | 0,74 | 92 |

AUC of ROC, Sensitivity, Specificity, Effect Size (Hedges m) for RP, SD & LHON patients computed against healthy participants. **Top** : AUC of ROC computed for both eyes with the spectral power of the 9 FOIs of 2 trials.
**Bottom**: AUC of ROC computed for both eyes. with the spectral power of the 9 FOIs of 2 trials and the slopes of a linear fit applied to the blink corrected pupillary traces. The number of observations is indicated in the last column.

Supplementary Table 5

| Test-Retest Run 1 vs. Run 2 RE & LE | $r^2$ | p | r |
|---|---|---|---|
| SD | 0,54 | <0,0001 | 0,73 |
| RP | 0,57 | <0,0001 | 0,75 |
| LHON | 0,7 | <0,0001 | 0,84 |
| HP | 0,72 | <0,0001 | 0,85 |

Test-Retest Pearson4s correlations for each group

| Correlation between RNFL & Spectral Power | $r^2$ | p | r |
|---|---|---|---|
| SD | 0,31 | 0,98 | 0,56 |
| RP | 0,206 | 0,52 | 0,45 |
| LHON | 0,154 | 0,07 | 0,39 |
| HP | 0,57 | 1E-05 | 0,75 |

Correlations between RNFL & MFPT Spectral Power for each group

Classification scores for the Quadrant stimuli

**Right Eye**

| RE x 2 Quadrants | AUC | Ss | Sp |
|---|---|---|---|
| SD | 0,96 | 0,92 | 0,89 |
| RP | 1 | 1 | 1 |
| LHON | 0,9 | 0,82 | 0,93 |
| **RE Quadrant 1** | **AUC** | **Ss** | **Sp** |
| SD | 1 | 1 | 1 |
| RP | 1 | 1 | 1 |
| LHON | 1 | 1 | 1 |
| **RE Quadrant 2** | **AUC** | **Ss** | **Sp** |
| SD | 1 | 1 | 1 |
| RP | 1 | 1 | 1 |
| LHON | 1 | 1 | 1 |

**Left Eye**

| LE x 2 Quadrants | AUC | Ss | Sp | N |
|---|---|---|---|---|
| SD | 0,93 | 0,86 | 0,89 | 56 |
| RP | 1 | 1 | 1 | 54 |
| LHON | 0,9 | 0,88 | 0,86 | 46 |
| **LE Quadrant 1** | **AUC** | **Ss** | **Sp** | **N** |
| SD | 1 | 1 | 1 | 28 |
| RP | 1 | 1 | 1 | 28 |
| LHON | 1 | 1 | 1 | 23 |
| **LE Quadrant 2** | **AUC** | **Ss** | **Sp** | **N** |
| SD | 1 | 1 | 1 | 28 |
| RP | 1 | 1 | 1 | 28 |
| LHON | 1 | 1 | 1 | 23 |

AUC of ROC, Sensitivity and Specificity for the **Quadrant stimuli** for RP, SD & LHON patients. The number of observations is indicated in the last column. **Top**: Results for both quadrant stimuli (2 trials) ; **Middle**: Results for the first quadrant stimuli (1 trial) ; **Bottom**: Results for second quadrant stimuli (1 trials)
